# Inhaled Fluticasone for Outpatient Treatment of Covid-19: A Decentralized, Placebo-controlled, Randomized, Platform Clinical Trial

**DOI:** 10.1101/2022.07.12.22277548

**Authors:** Accelerating Covid-19 Therapeutic Interventions and Vaccines (ACTIV)-6 Study Group, Susanna Naggie

**Author notes:** ACTIV-6 Study Group Members listed at the end of the manuscript.

## Abstract

**Background:** The effectiveness of inhaled corticosteroids to shorten time to symptom resolution or prevent hospitalization or death among outpatients with mild-to-moderate coronavirus 2019 (Covid-19) is unclear.

**Methods:** ACTIV-6 is an ongoing, decentralized, double-blind, randomized, placebo-controlled platform trial testing repurposed medications in outpatients with confirmed SARS-CoV-2 infection. Non-hospitalized adults aged ≥30 years, experiencing ≥2 symptoms of acute infection for ≤7 days were randomized to inhaled fluticasone furoate 200 μg once daily for 14 days or placebo. The primary outcome was time to sustained recovery, defined as the third of 3 consecutive days without symptoms. Secondary outcomes included composites of hospitalization or death with or without urgent care or emergency department visit by day 28.

**Results:** Of those eligible for the fluticasone arm, 656 were randomized to and received inhaled fluticasone; 621 received concurrent placebo. There was no evidence of improvement in time to recovery with fluticasone compared with placebo (hazard ratio [HR] 1.01, 95% credible interval [CrI] 0.91–1.12; posterior probability for benefit [HR>1]=0.56). Twenty-four participants (3.7%) in the fluticasone arm had urgent care or emergency department visits or were hospitalized compared with 13 (2.1%) in the pooled, concurrent placebo arm (HR 1.9, 95% CrI 0.8–3.5; posterior probability for benefit [HR<1]=0.03). Three participants in each arm were hospitalized, and no deaths occurred. Adverse events were uncommon in both arms.

**Conclusions:** Treatment with inhaled fluticasone furoate for 14 days did not result in improved time to recovery among outpatients with Covid-19 in the United States during the delta and omicron variant surges.

**Trial Registration:** ClinicalTrials.gov (NCT04885530).

## INTRODUCTION

As the severe acute respiratory syndrome coronavirus 2 (SARS-CoV-2) pandemic continues, there is an ongoing need for early therapies to prevent progression to severe coronavirus disease 2019 (Covid-19). Novel oral antivirals are being used to an increasing degree in high-income countries,^1–3^ with a clear clinical benefit in unvaccinated persons.^2^ Yet antivirals are unavailable in most low- and middle-income country settings and vaccination rates are variable. Thus, effective therapies are still needed for those with symptomatic infection to hasten clinical recovery.

Numerous repurposed drugs have been investigated for potential therapeutic effects.^4–7^ With dexamethasone being effective among hospitalized patients with hypoxia,^8^ inhaled corticosteroids have been tested as a possible early outpatient therapy, with inconsistent results.^9–13^ Open-label randomized trials with inhaled budesonide given 800 μg twice daily for 14 days showed benefits for faster time to recovery and strong trends in decreased hospitalizations or deaths.^12,13^ Conversely, randomized trials with inhaled ciclesonide (640 μg/day), 2 of which were double-blind, showed no change in symptom duration with variable decreases or possible increases in healthcare utilization.^9–11^ Trial sizes varied from 146 to 2530 participants, with both the largest and smallest trial reporting a statistically significant benefit over placebo.^12,13^ All 5 of these trials of inhaled corticosteroids were conducted among predominantly unvaccinated persons.^9–13^ The conflicting results led to regulatory authorities and guideline committees not recommending inhaled corticosteroid therapy as early treatment for Covid-19.

We sought to investigate inhaled fluticasone furoate in a double-blind, randomized, placebo-controlled, platform trial investigating repurposed drugs for non-hospitalized persons with mild-to-moderate Covid-19. Fluticasone propionate has approximately 4-fold more relative systemic steroid potency than budesonide,^14^ and fluticasone furoate has greater receptor affinity than the propionate ester allowing for once daily dosing.^15^ With enrollment to date largely representing the delta and omicron (BA.1.1) variants and in the post-vaccine era, we report here on the efficacy of inhaled fluticasone furoate at a dose of 200 µg daily for 14 days as compared with placebo for the treatment of early mild-to-moderate Covid-19.

## METHODS

### Trial Design and Oversight

Accelerating Covid-19 Therapeutic Interventions and Vaccines (ACTIV)-6 (NCT04885530) is a double-blind, randomized, placebo-controlled platform protocol conducted using a decentralized approach. ACTIV-6 enrolls outpatients with mild-to-moderate Covid-19 with a confirmed positive polymerase chain reaction (PCR) or antigen test for SARS-CoV-2 infection, including home-based testing.

The protocol was approved by each site’s institutional review board. Informed consent was obtained from each participant either via written consent or an e-consent process. An independent data monitoring committee oversaw participant safety and trial performance.

### Participants

The overall platform trial opened to recruitment on June 23, 2021 and is ongoing. Participants were enrolled into the inhaled fluticasone furoate arm or matched placebo inhaler or contributing placebo from August 10, 2021 through February 12, 2022 at 93 sites in the United States. Participants were either recruited by trial sites or self-identified online or by contacting a central study telephone hotline. Participants without a local site were managed centrally.

Sites verified eligibility criteria including age ≥30 years, confirmed SARS-CoV-2 infection ≤10 days, and experiencing ≥2 Covid-19 symptoms for ≤7 days from enrollment. Symptoms included the following: fatigue, dyspnea, fever, cough, nausea, vomiting, diarrhea, body aches, chills, headache, sore throat, nasal symptoms, and new loss of sense of taste or smell. Exclusion criteria included hospitalization, known allergy or contraindication including prohibited concomitant medications, or study drug use within 14 days of enrollment (see **Supplementary Appendix**). Fluticasone-specific self-reported exclusion criteria were pregnancy, breastfeeding, milk protein hypersensitivity, or use within ≤30 days of inhaled or systemic corticosteroids. Vaccination was not an exclusion. Standard of care Covid-19 therapies available under US Food and Drug Administration (FDA)-approval or emergency use authorization were allowable.

### Randomization and Interventions

Within the platform trial, study drugs could be added or removed according to adaptive design and/or emerging evidence. Participants were randomized to 1 of the study drugs actively enrolling at the time of randomization, which included ivermectin 400 μg/kg/day or fluvoxamine 50 mg twice daily during the period the fluticasone furoate arm was open. As multiple study drugs were available, randomization occurred based on appropriateness of each drug for the participant as determined by the eligibility criteria. Participants could choose to opt out of specific arms during the consent process.

ACTIV-6 was designed to share information from participants randomized to receive placebo. At the first step of randomization, participants were assigned to receive either placebo or intervention in the ratio 1:*m*, where *m* is the number of study drug arms for which participants were eligible. Subsequently, participants were randomized among the *m* study arms with equal probability. Participants randomized to receive placebo contribute to all of the study arms, regardless of type of matched-placebo comparator, for which eligibility was met at the time of randomization.

Via direct home delivery from a central pharmacy participants received an inhaler with a 14-day supply of either fluticasone furoate provided as a dry powder in a foil blister strip or identical matched placebo foil blister strip, provided by the manufacturer (GSK, Brentford, United Kingdom). Using the inhaler, participants self-administered 200 µg (1 blister) of fluticasone furoate or matching-placebo once daily for 14 days. Other concurrent placebos included ivermectin-matched placebo for 3 days or fluvoxamine-matched placebo for 14 days.

### Outcome Measures

The primary measure of effectiveness was time to recovery, defined as the third of 3 consecutive days without symptoms. This was selected *a priori* from among the 2 co-primary endpoints that remain available to other platform study drugs. The key secondary outcome is hospitalization or death by day 28. Other secondary outcomes included time unwell with ongoing symptoms; Covid-19 Clinical Progression Scale on days 7, 14, and 28; mortality through day 28; and urgent care visit, emergency department visit, or hospitalization through day 28.

### Trial Procedures

ACTIV-6 is a decentralized trial; thus, all study visits are planned as remote. Screening and eligibility confirmation are participant-reported and confirmed by trial sites. A positive SARS-CoV-2 test result was verified by sites prior to randomization. At the screening visit, participants reported demographic information, eligibility criteria, medical history, concomitant medications, symptom reporting, and quality of life questionnaires.

A central investigational pharmacy distributed study drug to residential addresses provided by participants, and shipping and delivery were tracked. Participants must have received study drug to be included in the analysis. Receipt of study drug was defined as day 1.

Participants were asked to complete assessments and report safety events daily through the first 14 days of study. From days 15–28, participants continued to report if they had symptoms until they had experienced 3 consecutive days without symptoms. Follow-up visits occurred at day 28 and day 90. At each study assessment, participants self-reported symptoms and severity, health care visits, and any new medications.

The daily and follow-up assessments were monitored, and sites were actively notified of events requiring review, including for reporting that meets criteria for serious adverse events (SAE) or unanticipated adverse device effects for the inhaler. In addition, participants were invited during assessments to request contact from the study team or to report any unusual circumstances. Failure to complete daily assessments also triggered a review for any possible SAEs. A missed assessment on the day after receiving the first dose of study medication (day 2) or any day of missed assessments up to day 14 prompted a notification to the site to contact the participant. All participants were instructed to self-report concerns either via an online event reporting system, by calling the site, or by calling a 24-hour hotline.

Events of special interest and SAEs were extracted by site investigators from the participants’ medical record if participants sought medical care or hospitalization occurred. Any medical events occurring after enrollment but that started before the receipt of study drug (day 1) were not considered adverse events or endpoints.

### Interim Analysis

Due to extremely rapid enrollment related to the omicron variant surge, 2000 participants were enrolled in ACTIV-6 from December 15, 2021 to February 1, 2022. This resulted in the rapid and full accrual of the fluticasone arm before the first planned interim analysis by the independent data monitoring committee.

### Statistical Analysis Plan

The ACTIV-6 trial is designed to be analyzed using a Bayesian approach. Decision thresholds were set to balance overall power with control of the Type I error rate in the context of a study drug-specific goal. An estimated sample size of approximately 1200 participants was expected to be sufficient to conclude whether there is meaningful evidence of clinical benefit for inhaled fluticasone for the primary endpoint.

As a platform trial, the primary analysis is implemented separately for each study drug, where the placebo group consists of concurrently randomized participants who met enrollment criteria for that study drug, including consent to be randomized to fluticasone. A modified intention-to-treat (mITT) approach was specified for primary analyses, including all participants who received study drug. From other remote trials,^4,7^ we recognized that medication delivery (placebo or study drug) may not always occur (e.g., failure of delivery, participant withdrawal, or intervening hospitalization prior to receipt). This resulted in exclusion of the participant for the mITT primary analysis. All available data were utilized to compare fluticasone furoate versus concurrent placebo control, regardless of post-randomization adherence. The safety population included those participants in the mITT population who reported taking at least 1 dose of study drug or matching placebo.

Heterogeneity in treatment effect was assessed for preselected subgroups and for the following covariates: age, sex, duration of symptoms, body mass index (BMI), symptom severity, calendar time (corresponding to predominant SARS-CoV-2 variant), and vaccination status.

## RESULTS

Among the 3750 participants in the platform trial, 1277 were eligible for this study arm and randomized to inhaled fluticasone furoate 200 µg/day (n=656) or placebo (n=621) (**Figure 1**). Of participants receiving placebo, 350 (56%) received matching placebo and 271 (44%) contributed from the concurrent placebo group.

**Figure 1.**
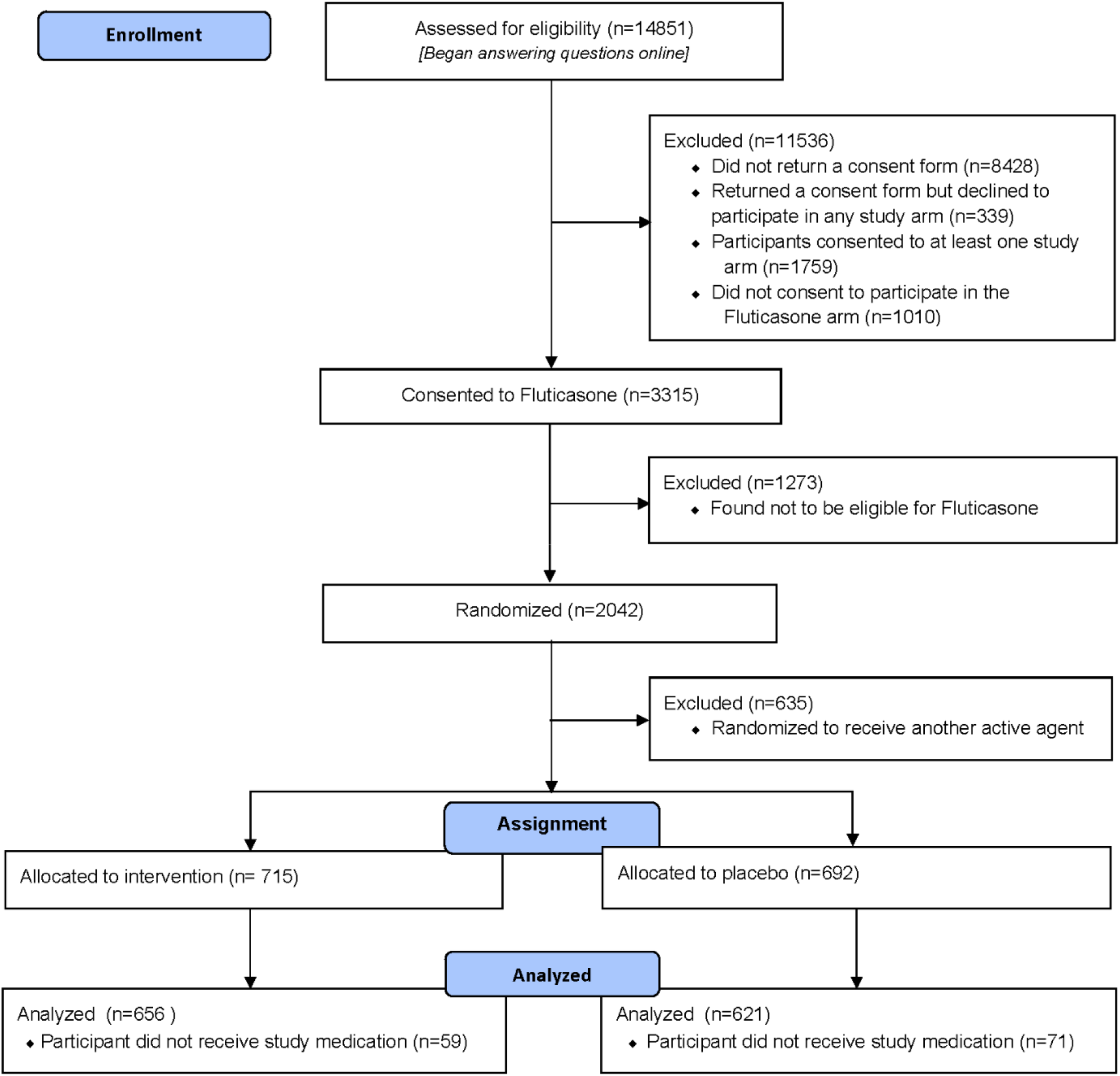
CONSORT diagram

The mean age of the participants was 47 years (SD ±12) and 39% were ≥50 years of age (**Table 1**). The population was 63% female, 80% identified as White, 7% Black/African American, 5.0% Asian, and 12.6% Latino/Hispanic ethnicity. Although not required for enrollment, co-morbidities were prevalent, including BMI >30 kg/m^2^ (39%), diabetes (9.5%), hypertension (25%), and asthma (13%). Vaccination was common, with 65% reporting at least 2 doses of a vaccine series. The median time from symptom onset to receipt of study drug was 5 days (IQR, 4 to 7). Baseline symptom prevalence and severity are described in **Table S1**. Although allowable per protocol, therapeutics available under FDA approval or authorization were uncommonly used (remdesivir 0.1%, monoclonal antibody 2.4%, ritonavir-boosted nirmatrelvir 0.1%) (**Table S2**).

**Table 1.** Baseline demographics of inhaled fluticasone furoate vs. concurrent placebo

| Variable | Inhaled Fluticasone<br>(n=656) | Placebo<br>(n=621) | Total<br>(N=1277) |
| --- | --- | --- | --- |
| Age, median (IQR), years | 45 (37, 55) | 46 (38, 56) | 45 (37, 55) |
| Age <50 years | 405 (61.7) | 370 (59.6) | 775 (60.7) |
| Female sex | 431 (65.7) | 376 (60.5) | 807 (63.2) |
| Race* |  |  |  |
| Black or African American | 47 (7.2) | 44 (7.1) | 91 (7.1) |
| White | 523 (79.7) | 500 (80.5) | 1023 (80.1) |
| Ethnicity: Latino, no./No. (%) | 78/655 (11.9) | 83/620 (13.4) | 161/1275 (12.6) |
| United States Region |  |  |  |
| Midwest | 141 (21.5) | 115 (18.5) | 256 (20.0) |
| Northeast | 52 (7.9) | 56 (9.0) | 108 (8.5) |
| South | 368 (56.1) | 371 (59.7) | 739 (57.9) |
| West | 95 (14.5) | 79 (12.7) | 174 (13.6) |
| Call center | 96 (14.6) | 97 (15.6) | 193 (15.1) |
| BMI, median (IQR), kg/m<sup>2</sup> | 28.1 (24.4, 33.6) | 28.1 (24.6, 32.9) | 28.1 (24.4, 33.4) |
| BMI >30 kg/m<sup>2</sup>, no./No. (%) | 260 (39.6) | 239/620 (38.5) | 499/1276 (39.1) |
| Heart disease, no./No. (%) | 25/640 (3.9) | 33/606 (5.4) | 58/1246 (4.65) |
| Diabetes, no./No. (%) | 56/640 (8.8) | 65/606 (10.7) | 121/1246 (9.71) |
| High blood pressure, no./No. (%) | 156/640 (24.4) | 169/606 (27.9) | 325/1246 (26.08) |
| COPD, no./No. (%) | 7/640 (1.1) | 11/606 (1.8) | 18/1246 (1.44) |
| Asthma, no./No. (%) | 76/640 (11.9) | 86/606 (14.2) | 162/1246 (13.00) |
| Chronic kidney disease, no./No. (%) | 6/640 (0.9) | 4/606 (0.7) | 10/1246 (0.80) |
| Smoker, past year, No./total (%) | 83/640 (13.0) | 72/606 (11.9) | 155/1246 (12.44) |
| Malignant cancer | 20 (3.1) | 23 (3.7) | 43 (3.37) |
| Vaccine status |  |  |  |
| Not vaccinated | 221 (33.7) | 211 (34.0) | 432 (33.8) |
| Vaccinated (1 dose) | 7 (1.1) | 11 (1.8) | 18 (1.4) |
| Vaccinated (2+ doses) | 428 (65.2) | 399 (64.3) | 827 (64.8) |
| Days between symptom onset and | 6 (4, 7) | 5 (4, 7) | 5 (4, 7) |

| **Variable** | **Inhaled Fluticasone**<br>**(n=656)** | **Placebo**<br>**(n=621)** | **Total**<br>**(N=1277)** |
| --- | --- | --- | --- |
| receipt of drug, median (IQR) |  |  |  |

Values are no. (%), unless otherwise noted.
\*Participants may have selected any combination of the following race descriptors: American Indian or Alaska Native; Asian; Black, African American, or African; Middle Eastern or North African; Native Hawaiian or Other Pacific Islander; White; None of the above; Prefer not to answer.
BMI indicates body mass index; COPD, chronic obstructive pulmonary disease; IQR, interquartile range.
Table S1 provides symptom distribution on study day 1 at time of receipt of study drug.

### Primary and Secondary Outcomes

In the mITT population, we observed no evidence of a difference in the primary outcome of time to recovery between the inhaled fluticasone furoate and placebo arms (hazard ratio [HR] 1.01, 95% credible interval ^1^ 0.91–1.12; posterior probability [HR>1]=0.56) (**Table 2, Figure 2, Figure S1**). There was also no difference in secondary outcomes measured through day 28 (**Figures S2-S5**). There were no deaths and hospitalizations were uncommon, occurring in only 0.5% (3/656) of the fluticasone furoate arm as compared with 0.5% (3/621) of the placebo arm. The composite secondary outcome of urgent care and emergency department visits, hospitalizations, or death had numerically higher events in the fluticasone furoate arm (24/656 [3.7%]) as compared with the placebo arm (13/621 [2.1%]) (**Table 2**). The Covid-19 Clinical Progression Scale (**Supplementary Appendix)** at days 7, 14, and 28 also did not meet prespecified thresholds for demonstrating any beneficial treatment effect. For example, by day 7, 86.4% (567/656) of the fluticasone furoate arm and 87.9% (546/621) of the placebo arm were not hospitalized and did not have any limitation of activities (**Table S3**).

**Table 2.**
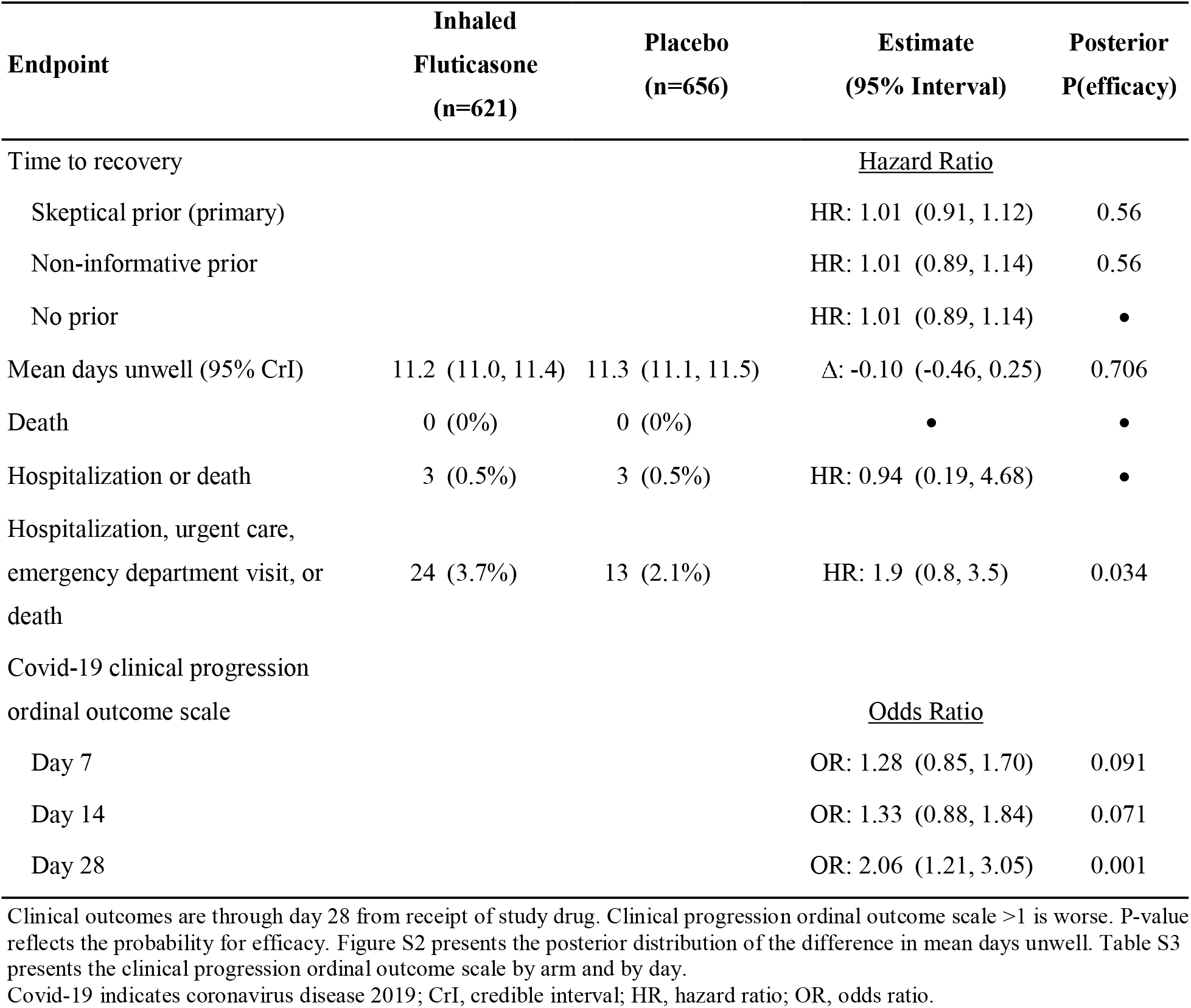
Primary and secondary clinical outcomes through day 28 of inhaled fluticasone furoate vs. concurrent placebo

**Figure 2.**
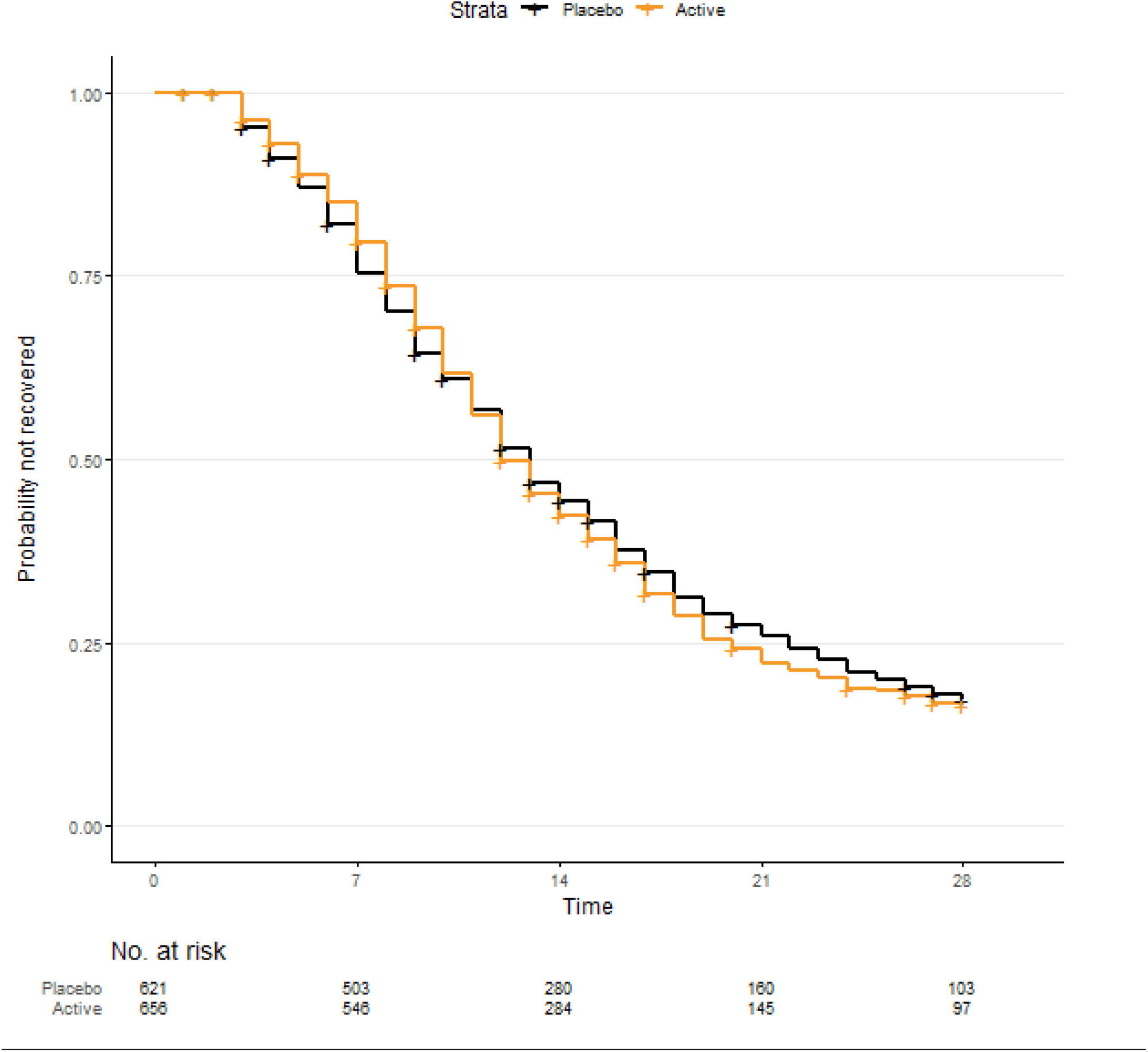
Time to sustained recovery from Covid-19 with inhaled fluticasone furoate versus concurrent placebo Kaplan-Meier curve for time-to-recovery primary endpoint. Time to sustained recovery was the number of days between the receipt of study drug and achieving 3 consecutive days without COVID-19 symptoms, as affirmatively reported by study participants. Participants who died, by definition, did not recover regardless of reported symptom freedom. Time to recovery was administratively censored at 28 days. The posterior probability was 0.56 for a faster recovery with inhaled fluticasone furoate versus placebo. **Figure S1** presents the posterior distribution of treatment effect. **Figure S4** presents sensitivity analyses for alternative methods of handling missing daily symptom data.

### Heterogeneity of Treatment Effect Analyses

There was no evidence of a treatment effect for time to recovery with fluticasone as compared with placebo for timing of symptom onset, severity of symptoms, BMI, age, sex, or calendar time. Differential treatment effects were noted for vaccination status (interaction P=0.02). Among the 845 participants who were vaccinated, those in the fluticasone arm trended towards faster recovery (HR 1.10, 95% confidence interval [CI] 0.95–1.28) than those in the placebo arm. However, among unvaccinated participants, those in the fluticasone arm trended towards longer time to recovery (HR 0.83, 95% CI 0.66–1.03) than those in the placebo arm, suggesting there may be heterogeneity in treatment response (**Figure 3**).

**Figure 3.**
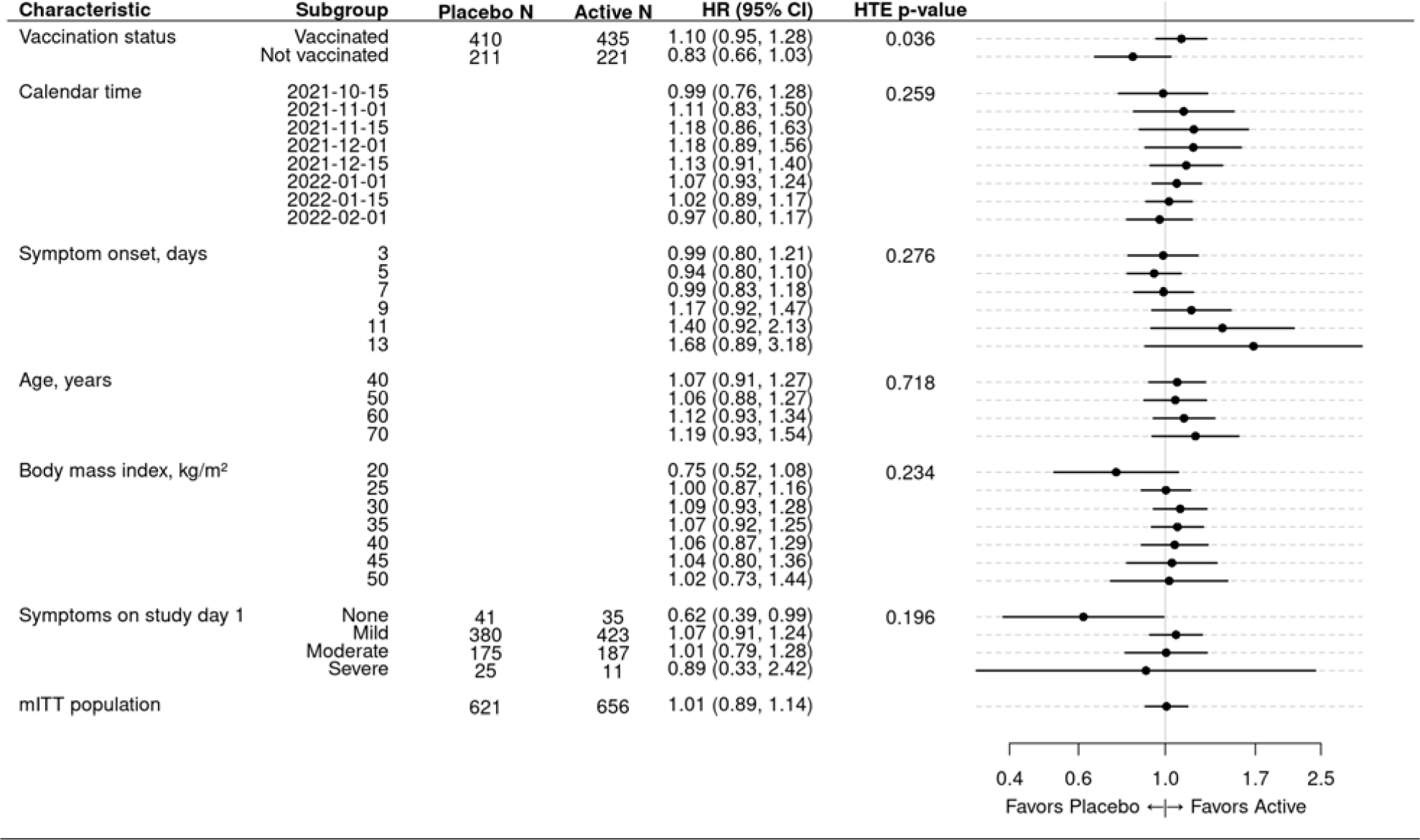
Heterogeneity of treatment effect between inhaled fluticasone furoate and concurrent placebo for time to recovery A hazard ratio greater than 1.0 indicates a faster time to recovery. Study day 1 was the day of starting the study medication. The ‘mITT population’ reflects a modified intent-to-treat analysis of participants randomized who enrolled within 7 days of symptom onset and received study drug. Covariate-adjusted and model-based estimates of the treatment effect for selected subgroups. For each characteristic, a proportional hazards regression model was constructed using the same covariates as the primary endpoint model plus additional interaction terms between treatment assignment and the characteristic of interest. For example, the interaction of vaccination status and treatment assignment was added to the primary endpoint regression model to calculate a treatment effect for the vaccinated and unvaccinated subgroups. To allow the possibility of non-linear trends along continuous characteristics, such as age or calendar time, the additional terms were interactions between treatment assignment and restricted cubic splines. Because the primary endpoint model did not include body mass index (BMI), the restricted cubic spline terms for BMI were also added to the model (sometimes called main effects) in addition to the interaction terms. Because the primary endpoint model only included a single linear term for symptom onset, the nonlinear terms of the restricted cubic spline were also added to the model in addition to the interaction terms. The hazard ratios and 95% confidence intervals were calculated from asymptotic, model-based contrasts. The hazard ratio for the full study population was generated from the primary endpoint model without a prior statistical inference of the possible effect.

### Safety

Among participants who reported taking study drug at least once, AEs were uncommon and similar in both arms (2% [13/640] with fluticasone vs. 2.5% [16/605] with placebo) (**Table S4**).

## DISCUSSION

We did not find beneficial treatment effects with inhaled fluticasone furoate 200 µg/day in this large randomized, double-blind trial that enrolled 1277 participants. The lack of treatment effect was consistent for the primary outcome of time to recovery with symptom resolution. Although no deaths occurred and hospitalizations were rare and similar in both groups, the number of acute care visits was higher with fluticasone than with placebo. Overall, the lack of treatment effect and the possible increased acute healthcare visits observed with inhaled fluticasone furoate (3.2%) over placebo (1.6%) makes this an unfavorable Covid-19 therapy.

There are numerous conflicting trials for use of inhaled steroids, and some of the differences may be due to variability in trial populations or to small or moderate sample sizes.^9–12^ In comparison with the largest published trial to date, the United Kingdom PRINCIPLE trial,^13^ ACTIV-6 has some similarities and differences. The timing of treatment was similar, as the PRINCIPLE trial initiated inhaled budesonide at a median of 6 days (IQR 4 to 9) from symptom onset. In theory, the relative steroid daily dose given in the PRINCIPLE trial was similar to fluticasone furoate 200 μg/day as fluticasone is approximately 4 times more potent than budesonide 800 μg twice daily and the fluticasone furoate formulation has a longer half-life.^14,15^

However, these are different steroids, and there could be a budesonide-specific effect. Two other significant differences include the methodology and study populations. ACTIV-6 utilized a double-blind, placebo-controlled trial design whereas PRINCIPLE was a pragmatic open-label trial, where the control group received usual care and not placebo.^13^ Whether a placebo effect could account for the 2.9 days faster time to recovery in PRINCIPLE seems unlikely, yet ACTIV-6 observed no improvement in time to recovery. Lastly, these trials enrolled different populations. PRINCIPLE enrolled unvaccinated persons >65 years old or >50 with a comorbidity (overall mean age of 65 years), whereas ACTIV-6 enrolled any adult ≥30 years (mean age 47 years) of whom 65% were vaccinated. The differences in age and vaccination status could result in a lower symptom severity in the patient population and less opportunity for treatment effects. In ACTIV-6, those with advanced age (>70 years) had the most favorable benefit (HR 1.19; 95% CrI, 0.93–1.54) for faster symptom resolution, but PRINCIPLE did not observe any differential effect by age.^13^ Similarly, in ACTIV-6 those who reported completing a vaccine series had a trend toward favorable benefit (HR 1.10; 95% CrI, 0.95–1.28), but PRINCIPLE did not include vaccinated people.

ACTIV-6 has several strengths. As a nationwide trial in the United States, ACTIV-6 is a generalizable trial for all adults aged ≥30 years old with Covid-19. This trial enrolled rapidly during the delta and omicron variant surges and included vaccinated patients, thus remaining a highly relevant population for the present times. ACTIV-6 also has limitations. Due to the broadly inclusive study population, few clinical events occurred, limiting the power to study the treatment effect on clinical outcomes like hospitalization. Due to the remote nature of the trial, the time from symptom onset to receipt of study drug was 6 days, which is later than the recommend start of antiviral medicines targeting ≤5 days.^2,3^ However, we did not observe any significant interaction with respect to the duration of symptoms when starting.

The ACTIV-6 trial did not identify a clinically relevant treatment effect with inhaled fluticasone furoate 200 µg daily for 14 days as outpatient Covid-19 treatment. We did not observe any faster time to clinical recovery in the population studied, unlike prior open-label trials of inhaled steroids, nor impact on prevention of clinical progression.^12,13^

## Data Availability

Prior to deposition of the data in a public repository which will occur when the platform trial has concluded, persons may request the data by submitting a proposal. If the data can be used for the proposed purpose and it is consistent with the informed consent, then the data will be released under a Data Use Agreement.

## ACTIV-6 Study Group

David R. Boulware, Elizabeth Shenkman, Adrian F. Hernandez, Susanna Naggie, G. Michael Felker, Sybil Wilson, Allison DeLong, April Remaly, Rhonda Wilder, Christopher J. Lindsell, Thomas G. Stewart, Sean Collins, Sarah Dunsmore, Sam Bozzette, Gene Passamani, Stacey Adam, Florence Thicklin, Matthew William McCarthy, George Hanna, Adit Ginde, Mario Castro, Dushyantha Jayaweera, Mark Sulkowski, Nina Gentile, Kathleen McTigue, Kim Marschhauser

## Author Contributions

Author and collaborator contributions, including responsibility for decision to submit the manuscript, drafting of the initial manuscript, study conceptualization, investigation, data curation, formal analysis, study supervision, and review and editing of the manuscript, are provided in the Online Supplement. CL and TS directly accessed and verified the underlying study data. SN and AFH had access to all the study data and had final responsibility for the decision to submit the paper for publication.

