## Supplemental Appendix for "Inhaled Fluticasone for Outpatient Treatment of Covid-19: A Decentralized, Placebo-controlled, Randomized, Platform Clinical Trial"

### Supplementary Appendix

#### ACTIV-6 Executive Committee

Adrian F. Hernandez, Duke Clinical Research Institute (Clinical Coordinating Center PI)

Susanna Naggie, Duke Clinical Research Institute (Clinical Coordinating Center Co-PI)

G. Michael Felker, Duke Clinical Research Institute (Medical Monitor, blinded)

Sybil Wilson, Duke Clinical Research Institute

Allison DeLong, Duke Clinical Research Institute

April Remaly, Duke Clinical Research Institute

Rhonda Wilder, Duke Clinical Research Institute

Christopher J. Lindsell, Vanderbilt University Medical Center (Data Coordinating Center PI)

Thomas G. Stewart, Vanderbilt University Medical Center

Sean Collins, Vanderbilt University Medical Center

Sarah Dunsmore, National Center for Advancing Translational Sciences

Sam Bozzette, National Center for Advancing Translational Sciences

Gene Passamani, National Center for Advancing Translational Sciences

Stacey Adam, Foundation for the National Institutes of Health

David Boulware, University of Minnesota

Elizabeth Shenkman, University of Florida

Florence Thicklin, Stakeholder Advisory Committee

Matthew William McCarthy, Weill Cornell Medicine, Stakeholder Advisory Committee

George Hanna, Biomedical Advanced Research and Development Authority

#### ACTIV-6 Protocol Oversight Committee

David Boulware, University of Minnesota, POC Co-Chair

Elizabeth Shenkman, University of Florida, POC Co-Chair

Adrian F. Hernandez, Duke Clinical Research Institute (Clinical Coordinating Center PI)

Susanna Naggie, Duke Clinical Research Institute (Clinical Coordinating Center co-PI)

G. Michael Felker, Duke Clinical Research Institute (Medical Monitor, blinded)

Sybil Wilson, Duke Clinical Research Institute

Allison DeLong, Duke Clinical Research Institute

April Remaly, Duke Clinical Research Institute

Rhonda Wilder, Duke Clinical Research Institute

Christopher J. Lindsell, Vanderbilt University Medical Center (Data Coordinating Center PI)

Thomas G. Stewart, Vanderbilt University Medical Center

Sean Collins, Vanderbilt University Medical Center
Sarah Dunsmore, National Center for Advancing Translational Sciences
Sam Bozzette, National Center for Advancing Translational Sciences
Gene Passamani, National Center for Advancing Translational Sciences
Stacey Adam, Foundation for the National Institutes of Health
Florence Thicklin, Stakeholder Advisory Committee
Matthew William McCarthy, Weill Cornell Medicine, Stakeholder Advisory Committee
George Hanna, Biomedical Advanced Research and Development Authority
Adit Ginde, University of Colorado Denver – Anschutz
Mario Castro, University of Kansas Medical Center
Dushyantha Jayaweera, University of Miami
Mark Sulkowski, John Hopkins University
Nina Gentile, Lewis Katz School of Medicine at Temple University
Kathleen McTigue, University of Pittsburgh Medical Center
Kim Marschhauser, PCORI
Julia Garcia-Diaz, Ochsner Health

#### ACTIV-6 Clinical Trial Team

Adrian F. Hernandez, Duke Clinical Research Institute (Clinical Coordinating Center PI)
Susanna Naggie, Duke Clinical Research Institute (Clinical Coordinating Center Co-PI)
G. Michael Felker, Duke Clinical Research Institute (Medical Monitor, blinded)
Sybil Wilson, Duke Clinical Research Institute
Allison DeLong, Duke Clinical Research Institute
April Remaly, Duke Clinical Research Institute
Rhonda Wilder, Duke Clinical Research Institute
Christopher J. Lindsell, Vanderbilt University Medical Center (Data Coordinating Center PI)
Thomas G. Stewart, Vanderbilt University Medical Center

#### ACTIV-6 Independent Data Monitoring Committee

Voting Members:

Clyde Yancy, Northwestern University Feinberg School of Medicine (Chair)
Adaora Adimora, University of North Carolina, Chapel Hill (Vice-chair)
Susan Ellenberg, University of Pennsylvania

Kaleab Abebe, University of Pittsburgh
Arthur Kim, Massachusetts General Hospital
John D. Lantos, Children's Mercy Hospital
Jennifer Silvey-Cason, Participant representative

Statistical Data Analysis Center:
Frank Rockhold, Duke Clinical Research Institute (Lead faculty statistician)
Sean O'Brien, Duke Clinical Research Institute (Faculty statistician)
Frank Harrell, Vanderbilt University Medical Center (DCC-SDAC Faculty Liaison)
Zhen Huang, Duke Clinical Research Institute (Lead statistician)

#### ACTIV-6 Clinical Events Classification Committee

Renato Lopes, (CEC PI)
Faculty Reviewers: W. Schuyler Jones, Antonio Gutierrez, Robert Harrison, David Kong, Robert McGarrah
Fellow Reviewers: Michelle Kelsey, Konstantin Krychtiuk, Vishal Rao

#### ACTIV-6 Data Coordinating Center

David Aamodt, Vanderbilt University Medical Center
JaMario Ayers, Vanderbilt University Medical Center
Jess Collins, Vanderbilt University Medical Center
John Graves, Vanderbilt University Medical Center
James Grindstaff, Vanderbilt University Medical Center
Frank Harrell, Vanderbilt University Medical Center (DCC-SDAC Faculty Liaison)
Jessica Lai, Vanderbilt University Medical Center
Christopher J. Lindsell, Vanderbilt University Medical Center (Data Coordinating Center PI)
Itzel Lopez, Vanderbilt University Medical Center
Jessica Marlin, Vanderbilt University Medical Center
Alyssa Merkel, Vanderbilt University Medical Center
Sam Nwosu, Vanderbilt University Medical Center
Savannah Obregon, Vanderbilt University Medical Center
Dirk Orozco, Vanderbilt University Medical Center
Yoli Perez-Torres, Vanderbilt University Medical Center

Nelson Prato, Vanderbilt University Medical Center
Colleen Ratcliff, Vanderbilt University Medical Center
Max Rohde, Vanderbilt University Medical Center
Russell Rothman, Vanderbilt University Medical Center
Jana Shirey-Rice, Vanderbilt University Medical Center
Krista Vermillion, Vanderbilt University Medical Center
Thomas Stewart, Vanderbilt University Medical Center (Lead statistician)
Hsi-nien Tan, Vanderbilt University Medical Center
Seibert Tregoning, Vanderbilt University Medical Center
Meghan Vance, Vanderbilt University Medical Center
Amber Vongsamphanh, Vanderbilt University Medical Center
Maria Weir, Vanderbilt University Medical Center
Nicole Zaleski, Vanderbilt University Medical Center

**ACTIV-6 Primary Site Investigators / Study Coordinators**

**A New Start II, LLC:** William (Kelly) Vincent / Raina Vincent. **Advanced Medical Care, Ltd:** Ray Bianchi / Jen Premas. **AMRON Vitality and Wellness Center, LLC:** Diana Cordero-Loperena / Evelyn Rivera. **Ananda Medical Clinic:** Madhu Gupta / Greg Karawan & Carey Ziomek. **Arena Medical Group:** Joseph Arena / Sonaly DeAlmeida. **Assuta Family Medical Group APMC:** Soroush Ramin / Jaya Nataraj. **Boston Medical Center:** Michael Paasche-Orlow / Lori Henault & Katie Waite. **Bucks County Clinical Research:** David Miller / Ginger Brounce. **Christ the King Health Care, P.C.:** Constance George-Adebayo / Adeolu Adebayo. **Clinical Trials Center of Middle Tennessee:** Alex Slandzicki / Jessica Wallan. **Comprehensive Pain Management and Endocrinology:** Claudia Vogel / Sebastian Munoz. **David Kavtaradze MD, Inc.:** David Kavtaradze / Cassandra Watson. **David Singleton MD, PA:** David Singleton / Maria Rivon & Amanda Sevier. **Del Pilar Medical and Urgent Care:** Arnold Del Pilar / Amber Spangler. **DHR Health Institute for Research:** Sohail Rao / Luis Cantu. **Diabetes and Endocrinology Assoc. of Stark County:** Arvind Krishna / Kathy Evans, Tylene Falkner & Brandi Kerr. **Doctors Medical Group of Colorado Springs, P.C.:** Robert Spees / Mailyn Marta. **Duke University:** G. Michael Felker / Amanda Harrington. **Duke University Hospital:** Rowena Dolor / Madison Frazier, Lorraine Vergara & Jessica Wilson. **Elite Family Practice:** Valencia Burruss / Terri Hurst. **Emory University:** Igho Ofotokun / Laurel Bristow. **Essentia Health:** Rajesh Prabhu / Krystal Klicka & Amber Lightfeather. **Essential Medical Care, Inc.:** Vicki James / Marcella Rogers. **Express Family Clinic:** Pradeep Parihar / De'Ambra Torress. **Family Practice Doctors P.A.:** Chukwuemeka Oragwu / Ngozi Oguego. **First Care Medical Clinic:** Rajesh Pillai / Mustafa Juma. **Focus Clinical Research Solutions:** Ahab Gabriel / Emad Ghaly. **Franciscan Health Michigan City:** Dafer Al-Haddadin / Courtney Ramirez. **G&S Medical Associates, LLC:** Gammal Hassanien / Samah Ismail. **George Washington University Hospital:** Andrew Meltzer / Seamus Moran. **Geriatrics and Medical Associates:** Scott Brehaut / Angelina Roche. **GFC of Southeastern Michigan, PC:** Manisha Mehta / Nicole Koppinger. **Health Quality Primary Care:** Jose Baez / Ivone Pagan. **Highlands Medical Associates, P.A.:** Dallal Abdelsayed / Mina Aziz. **Hoag Memorial Hospital Presbyterian:** Philip Robinson / Julie Nguyen. **Hugo Medical Clinic:** Victoria Pardue / Llisa Hammons. **Innovation Clinical Trials Inc.:** Juan Ruiz-Unger / Susan Gonzalez & Lionel Reyes. **Jackson Memorial Hospital:** John Cienki / Gisselle Jimenez. **Jadestone Clinical Research, LLC:** Jonathan Cohen / Matthew Wong & Ying Yuan. **Jeremy W. Szeto, D.O., P.A.:** Jeremy Szeto. **Johns Hopkins Hospital:** Mark Sulkowski / Lauren Stelmash. **Lakeland Regional Medical Center:** Arch Amon / Daniel Haight. **Lamb Health, LLC:** Deryl Lamb / Amron Harper.

**Lice Source Services Plantation:** Nancy Pyram-Bernard / Arlen Quintero. **Lupus Foundation of Gainesville:** Eftim Adhami. **Maria Medical Center, PLLC:** Josette Maria / Diksha Paudel & Oksana Raymond. **Medical Specialists of Knoxville:** Jeffrey Summers / Tammy Turner. **Medical University of South Carolina:** Leslie Lenert / Sam Gallegos & Elizabeth Ann Szwast. **Mediversity Healthcare:** Ahsan Abdulghani / Pravin Vasoya. **Miller Family Practice, LLC:** Conrad Miller / Hawa Wiley. **North Shore University Health System/Evanston Hospital:** Nirav Shah / Tovah Klein. **Ochsner Clinic Foundation:** Julie Castex / Phillip Feliciano. **Olivo Wellness Medical Center:** Jacqueline Olivo / Marian Ghaly, Zainub Javed & Alexandra Nawrocki. **Pine Ridge Family Medicine Inc.:** Anthony Vecchiarelli / Nikki Vigil. **Premier Health:** Vijaya Cherukuri / Erica Burden. **Rapha Family Wellness:** Dawn Linn / Laura Fisher. **Raritan Bay Primary Care & Cardiology Associates:** Vijay Patel / Praksha Patel & Yuti Patel. **Romancare Health Services:** Leonard Ellison / Jeffrey Harrison. **Spinal Pain and Medical Rehab, PC:** Binod Shah / Sugata Shah. **Stanford University:** Upinder Singh / Julia Donahue & Yasmin Jazayeri. **Sunshine Walk In Clinic:** Anita Gupta / N Chandrasekar & Beth Moritz. **Tabitha B. Fortt, M.D., LLC:** Tabitha Fortt / Anisa Fortt. **Tallahassee Memorial Hospital:** Ingrid Jones-Ince / Alix McKee & Christy Schattinger. **Tampa General Hospital:** Jason Wilson / Brenda Farlow. **Temple University Hospital:** Nina Gentile / Lillian Finlaw. **Texas Health Physicians Group:** Randall Richwine & Tearani Williams / Penny Pazier & Lisa Carson. **Texas Tech University Health Sciences Center in El Paso:** Edward Michelson / Danielle Austin. **The Heart and Medical Center:** Sangeeta Khetpal / Tiffaney Cantrell, Drew Franklin & Karissa Marshall. **Trident Health Center:** Arvind Mahadevan / Madelyn Rosequist. **TriHealth, Inc:** Martin Gnoni / Crystal Daffner. **UF Health Precision Health Research:** Carla VandeWeerd / Mitchell Roberts. **University Diagnostics and Treatment Clinic:** Mark D'Andrea / Mina Aziz. **University Medical Center- New Orleans:** Stephen Lim / Wayne Swink. **University of Cincinnati:** Margaret Powers-Fletcher / Sylvere Mukunzi. **University of Florida Health:** Elizabeth Shenkman / Jamie Hensley & Brittney Manning. **University of Florida-JAX-ASCENT:** Carmen Isache / Jennifer Bowman, Angelique Callaghan-Brown & Taylor Scott. **University of Kansas – Wichita:** Tiffany Schwasinger-Schmidt / Ashlie Cornejo. **University of Miami:** Dushyantha Jayaweera / Maria Almanzar, Letty Ginsburg & Americo Hajaz. **University of Minnesota:** Carolyn Bramante. **University of Missouri – Columbia:** Matthew Robinson / Michelle Seithel. **University of Pittsburgh:** Akira Sekikawa / Emily Klawson. **University of Texas Health Science Center at Houston:** Luis Ostrosky / Virginia Umana. **University of Texas Health Science Center at San Antonio:** Thomas Patterson / Robin Tragus. **University of Virginia Health System:** Patrick Jackson / Caroline Hallowell & Heather Haughey. **Vaidya**

**MD PLLC:** Bhavna Vaidya-Tank / Cameron Gould. **Vanderbilt University Medical Center:** Parul Goyal / Carly Gatewood. **Wake Forest University Health Sciences:** John Williamson / Hannah Seagle. **Weill Cornell Medical College:** Matthew McCarthy / Elizabeth Salsgiver. **Well Pharma Medical Research:** Eddie Armas / Jhonsai Cheng & Priscilla Huerta.

**COVID-19 Ordinal Outcome Scale**

The COVID-19 outcomes for this trial are based on the World Health Organization's Ordinal Scale for Clinical Improvement and will be collected via the online system and from the medical record. The following outcomes will be assessed as part of the COVID Clinical Progression Scale:

0. No clinical or virological evidence of infection
1. No limitation of activities
2. Limitation of activities
3. Hospitalized, no oxygen therapy
4. Hospitalized, on oxygen by mask or nasal prongs
5. Hospitalized, on non-invasive ventilation or high-flow oxygen
6. Hospitalized, on intubation and mechanical ventilation
7. Hospitalized, on ventilation + additional organ support – pressors, RRT, ECMO
8. Death

**Table S1. Baseline prevalence and severity of symptoms on study day 1**

| Variable | Inhaled Fluticasone (n=656) | Placebo (n=621) | Total (N=1277) |
|---|---|---|---|
| Symptom burden on study day 1, No. (%) | | | |
| None | 35 (5.3%) | 41 (6.6%) | 76 (6.0%) |
| Mild | 423 (64.5%) | 380 (61.2%) | 803 (62.9%) |
| Moderate | 187 (28.5%) | 175 (28.2%) | 362 (28.3%) |
| Severe | 11 (1.7%) | 25 (4.0%) | 36 (2.8%) |
| Symptoms on study day 1 | 599 | 570 | 1169 |
| Fatigue | | | |
| None | 45 (7.5%) | 52 (9.1%) | 97 (8.3%) |
| Mild | 304 (50.8%) | 281 (49.3%) | 585 (50.0%) |
| Moderate | 211 (35.2%) | 200 (35.1%) | 411 (35.2%) |
| Severe | 39 (6.5%) | 37 (6.5%) | 76 (6.5%) |
| Dyspnea | | | |
| None | 317 (52.9%) | 313 (54.9%) | 630 (53.9%) |
| Mild | 223 (37.2%) | 192 (33.7%) | 415 (35.5%) |
| Moderate | 57 (9.5%) | 59 (10.4%) | 116 (9.9%) |
| Severe | 2 (0.3%) | 6 (1.1%) | 8 (0.7%) |
| Fever | | | |
| None | 466 (77.8%) | 434 (76.1%) | 900 (77.0%) |
| Mild | 107 (17.9%) | 106 (18.6%) | 213 (18.2%) |
| Moderate | 23 (3.8%) | 26 (4.6%) | 49 (4.2%) |
| Severe | 3 (0.5%) | 4 (0.7%) | 7 (0.6%) |
| Cough | | | |
| None | 84 (14.0%) | 78 (13.7%) | 162 (13.9%) |
| Mild | 334 (55.8%) | 305 (53.5%) | 639 (54.7%) |
| Moderate | 156 (26.0%) | 166 (29.1%) | 322 (27.5%) |
| Severe | 25 (4.2%) | 21 (3.7%) | 46 (3.9%) |
| Nausea | | | |
| None | 450 (75.1%) | 425 (74.6%) | 875 (74.9%) |
| Mild | 115 (19.2%) | 112 (19.6%) | 227 (19.4%) |
| Moderate | 28 (4.7%) | 25 (4.4%) | 53 (4.5%) |
| Severe | 6 (1.0%) | 8 (1.4%) | 14 (1.2%) |
| Vomiting | | | |
| None | 570 (95.2%) | 547 (96.0%) | 1117 (95.6%) |
| Mild | 24 (4.0%) | 16 (2.8%) | 40 (3.4%) |
| Moderate | 4 (0.7%) | 6 (1.1%) | 10 (0.9%) |
| Severe | 1 (0.2%) | 1 (0.2%) | 2 (0.2%) |
| Diarrhea | | | |
| None | 459 (76.6%) | 427 (74.9%) | 886 (75.8%) |
| Mild | 103 (17.2%) | 110 (19.3%) | 213 (18.2%) |
| Moderate | 30 (5.0%) | 28 (4.9%) | 58 (5.0%) |

| Variable | Inhaled Fluticasone (n=656) | Placebo (n=621) | Total (N=1277) |
|---|---|---|---|
| Severe | 7 (1.2%) | 5 (0.9%) | 12 (1.0%) |
| Body aches | | | |
| None | 207 (34.6%) | 184 (32.3%) | 391 (33.4%) |
| Mild | 241 (40.3%) | 250 (43.9%) | 491 (42.0%) |
| Moderate | 122 (20.4%) | 113 (19.8%) | 235 (20.1%) |
| Severe | 29 (4.8%) | 23 (4.0%) | 52 (4.4%) |
| Sore throat | | | |
| None | 286 (47.7%) | 263 (46.1%) | 549 (47.0%) |
| Mild | 237 (39.6%) | 218 (38.2%) | 455 (38.9%) |
| Moderate | 58 (9.7%) | 69 (12.1%) | 127 (10.9%) |
| Severe | 18 (3.0%) | 20 (3.5%) | 38 (3.3%) |
| Headache | | | |
| None | 242 (40.4%) | 224 (39.3%) | 466 (39.9%) |
| Mild | 246 (41.1%) | 234 (41.1%) | 480 (41.1%) |
| Moderate | 89 (14.9%) | 90 (15.8%) | 179 (15.3%) |
| Severe | 22 (3.7%) | 22 (3.9%) | 44 (3.8%) |
| Chills | | | |
| None | 427 (71.3%) | 403 (70.7%) | 830 (71.0%) |
| Mild | 125 (20.9%) | 123 (21.6%) | 248 (21.2%) |
| Moderate | 39 (6.5%) | 36 (6.3%) | 75 (6.4%) |
| Severe | 8 (1.3%) | 8 (1.4%) | 16 (1.4%) |
| Nasal symptoms | | | |
| None | 159 (26.5%) | 143 (25.1%) | 302 (25.8%) |
| Mild | 302 (50.4%) | 306 (53.7%) | 608 (52.0%) |
| Moderate | 119 (19.9%) | 99 (17.4%) | 218 (18.6%) |
| Severe | 19 (3.2%) | 22 (3.9%) | 41 (3.5%) |
| New loss of taste or smell | | | |
| None | 378 (63.1%) | 337 (59.1%) | 715 (61.2%) |
| Mild | 105 (17.5%) | 110 (19.3%) | 215 (18.4%) |
| Moderate | 57 (9.5%) | 63 (11.1%) | 120 (10.3%) |
| Severe | 59 (9.8%) | 60 (10.5%) | 119 (10.2%) |

Values are no. (%). Study day 1 was the day of receipt of study medication. Participants were required to have at least 2 symptoms at time of randomization.

#### Table S2. FDA-authorized therapeutics utilized by participants

| COVID-19 Therapeutic | Inhaled Fluticasone (n=656) | Placebo (n=621) |
|---|---|---|
| Remdesivir | 1 (0.2%) | 0 (0.0%) |
| Monoclonal antibodies | 17 (2.6%) | 13 (2.1%) |
| Nirmatrelvir/ritonavir | 0 (0.0%) | 1 (0.2%) |

Values are no. (%).

#### Table S3. Covid-19 clinical progression scale for inhaled fluticasone vs placebo

| Clinical Progression Scale | Day 7 | | Day 14 | | Day 28 | |
|---|---|---|---|---|---|---|
| | Fluticasone | Placebo | Fluticasone | Placebo | Fluticasone | Placebo |
| Not hospitalized, activity level not reported | 6.4% (42) | 4.35% (27) | 9.8% (64) | 6.6% (41) | 7.9% (52) | 4.7% (29) |
| Not hospitalized, no limitation of activities | 86.4% (567) | 87.9% (546) | 86.3% (566) | 88.1% (547) | 88.3% (579) | 92.4% (574) |
| Not hospitalized, limitation of activities | 6.4% (42) | 6.9% (43) | 2.9% (19) | 4.2% (26) | 1.7% (11) | 1.0% (6) |
| Hospitalized, no oxygen therapy | 0.0% (0) | 0.0% (0) | 0.0% (0) | 0.0% (0) | 0.0% (0) | 0.0% (0) |
| Hospitalized, on oxygen therapy | 0.0% (0) | 0.2% (1) | 0.0% (0) | 0.0% (0) | 0.0% (0) | 0.0% (0) |
| Hospitalized, on non-invasive ventilation or high-flow oxygen | 0.15% (1) | 0.0% (0) | 0.15% (1) | 0.0% (0) | 0.0% (0) | 0.0% (0) |
| Hospitalized, on intubation and mechanical ventilation | 0.0% (0) | 0.0% (0) | 0.0% (0) | 0.0% (0) | 0.0% (0) | 0.0% (0) |
| Hospitalized, on ventilation + additional organ support | 0.0% (0) | 0.0% (0) | 0.0% (0) | 0.0% (0) | 0.0% (0) | 0.0% (0) |
| Death | 0.0% (0) | 0.0% (0) | 0.0% (0) | 0.0% (0) | 0.0% (0) | 0.0% (0) |
| Missing Outcome | 0.6% (4) | 0.6% (4) | 0.9% (6) | 1.1% (7) | 2.1% (14) | 1.9% (12) |

Covid-19 clinical progression outcome scale; values are % (n).

#### Table S4. Adverse events experienced by participants

| Variable | Fluticasone, Not Taken (n=16) | Fluticasone, Taken (n=640) | Placebo, Not Taken (n=16) | Placebo, Taken (n=605) | Total (N=1277) |
|---|---|---|---|---|---|
| Experienced an adverse event | 0 (0%) | 13 (2.03%) | 0 (0%) | 16 (2.6%) | 29 (2.3%) |
| Experienced a serious adverse event | 0 (0%) | 3 (0.47%) | 0 (0%) | 6 (1.0%) | 9 (0.7%) |
| Serious adverse events | | | | | |
| Covid-19 pneumonia | | 1 | | 1 | 2 |
| Covid-19 pneumonia aggravated | | 2 | | 0 | 2 |
| Coronary vasospasm | | 0 | | 1 | 1 |
| Diplopia | | 0 | | 1 | 1 |
| Nausea and vomiting symptoms | | 0 | | 1 | 1 |
| Urinary tract infection | | 0 | | 1 | 1 |
| Adverse drug reaction | | 0 | | 1 | 1 |

Values are N (%). Note: “Taken” refers to the participants who reported taking (or planning to take) the study drug at least once. “Not taken” refers to the participants (if any) who did not report taking the study drug.
Covid-19 indicates coronavirus disease 2019.

**Figure S1. Posterior distribution of treatment effect hazard ratio for time to sustained recovery**

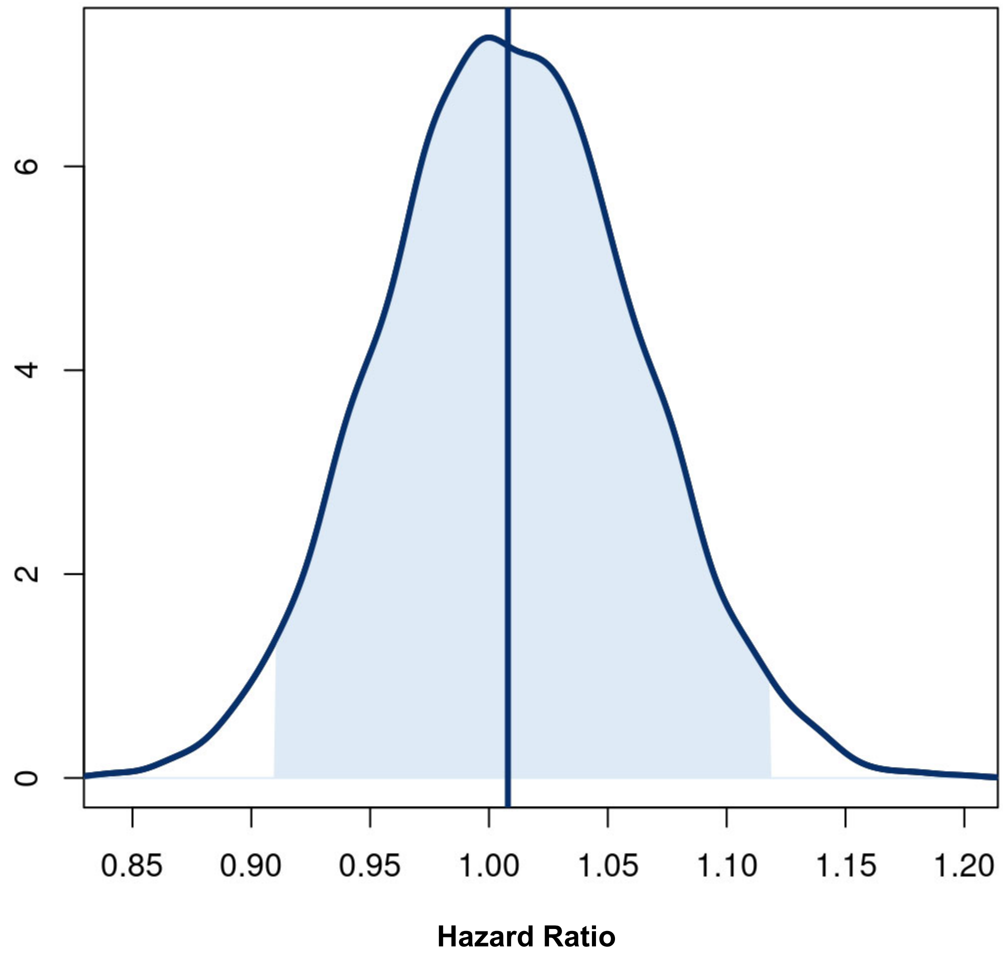

Posterior distribution for the treatment effect hazard ratio for the time to sustained recovery. The posterior was based on a covariate adjusted Cox proportional hazards regression with skeptical prior. The baseline hazard was a degree 5 M-spline function. Covariates in the model included age (as restricted cubic spline), sex, duration of symptoms, vaccine status, geographic region, origination from call center, calendar time (as restricted cubic spline), and symptom burden on the day of drug receipt.

The posterior probability was 0.56 for hazard ratio >1.0. Hazard ratios greater than one favored the active intervention for a faster time to recovery.

**Figure S2. Posterior distribution of the difference in mean time unwell**

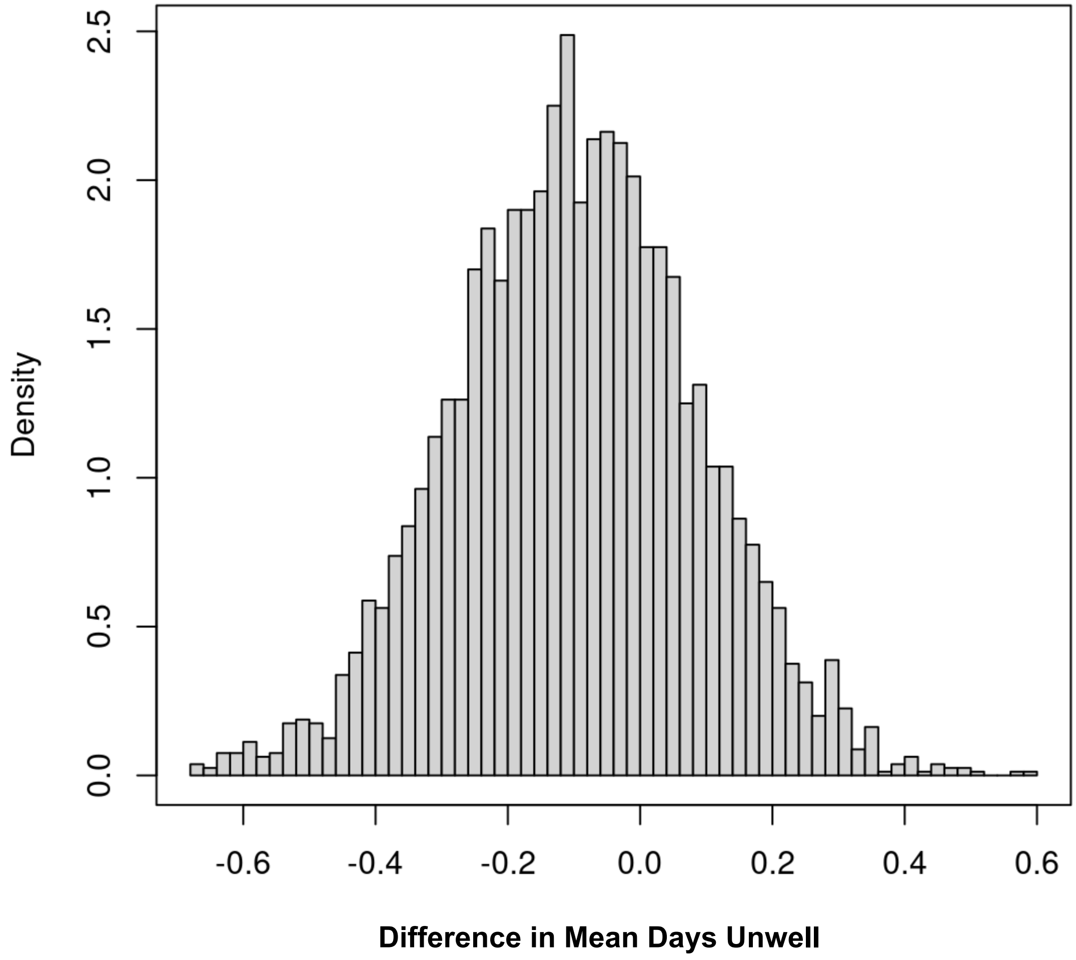

---

Posterior distribution of the difference in mean days unwell (Active-Placebo). Mean time unwell is a model-based estimate of the number of days with symptoms or hospitalized/deceased during the first 14 days of follow-up. Negative differences indicate that participants in the active arm were unwell shorter than participants in the placebo arm. The estimate of mean days unwell is calculated from a Bayesian, longitudinal, ordinal regression model with covariates age (as restricted cubic spline) and calendar time. The prior distribution was not informative, as the Bayesian statistical inference.

The difference in mean days unwell was -0.10 (95% CrI, -0.46, 0.25) equating to an average of 2.4 hours faster recovery with inhaled fluticasone (95% CrI, 11 hours faster to +6 hours longer). The posterior probability that the difference was larger than 1 day was less than 0.001 probability.

**Figure S3A. Time to composite endpoint of hospitalization, urgent care, emergency department visit, or death through day 28 for inhaled fluticasone furoate vs concurrent placebo**

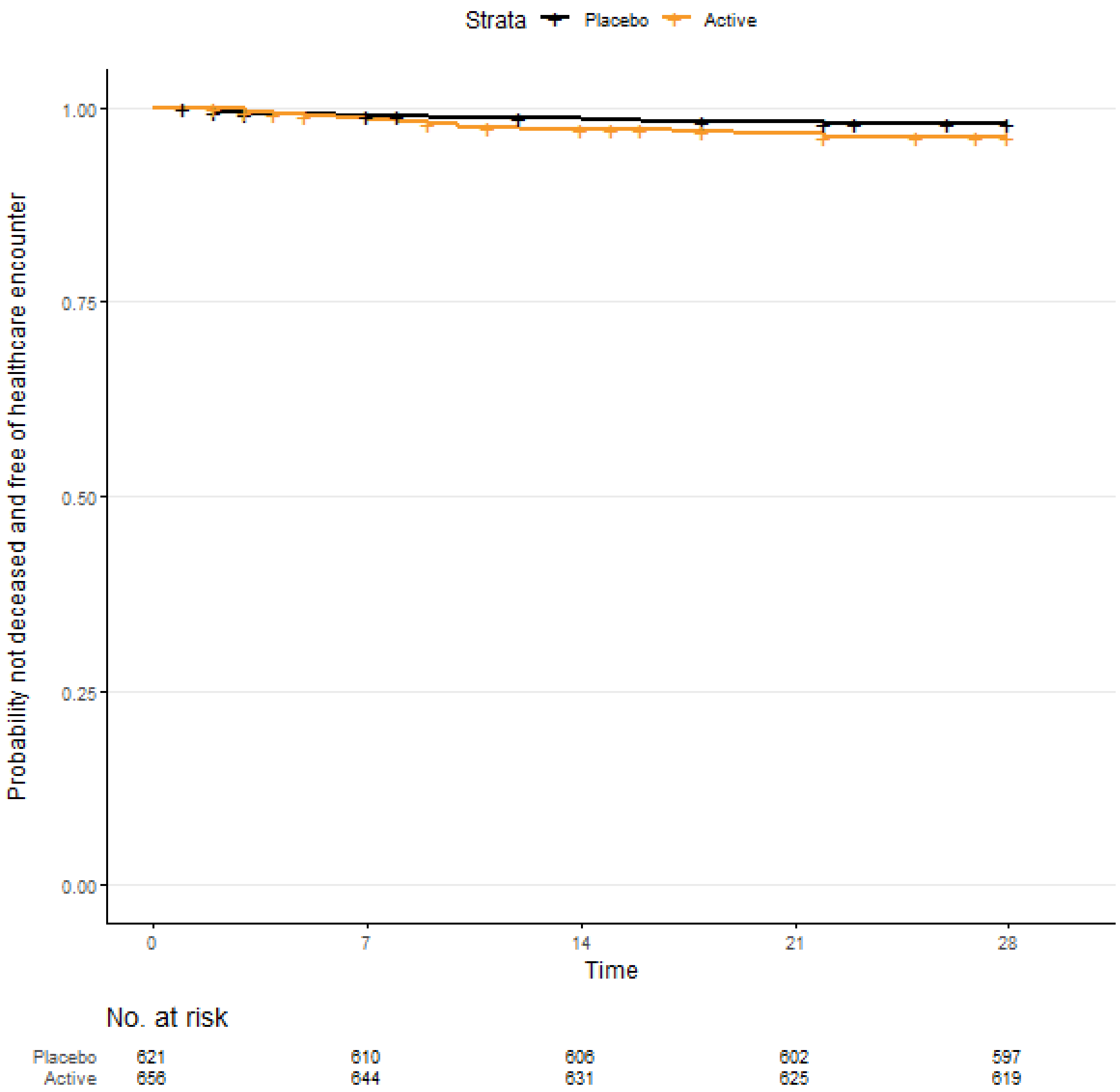

Kaplan-Meier curve for time-to-healthcare encounter/hospitalization/death endpoint. In the fluticasone furoate active group, 24 participants (3.7%) had a healthcare encounter of urgent care visit, emergency room visit, or hospitalization as compared with 13 (2.1%) in the pooled placebo group (Hazard Ratio 1.9, 95%CrI, 0.8-3.5, P(HR<1)=0.03). Overall, 13 fluticasone participants and 11 placebo participants were right-hand censored for lost to follow up. The Bayesian posterior distribution of treatment effect is presented in **Figure S3B**.

**Figure S3B. Posterior distributions of treatment effect hazard ratio for time to the composite endpoint of hospitalization, urgent care, emergency department visit, or death through day 28**

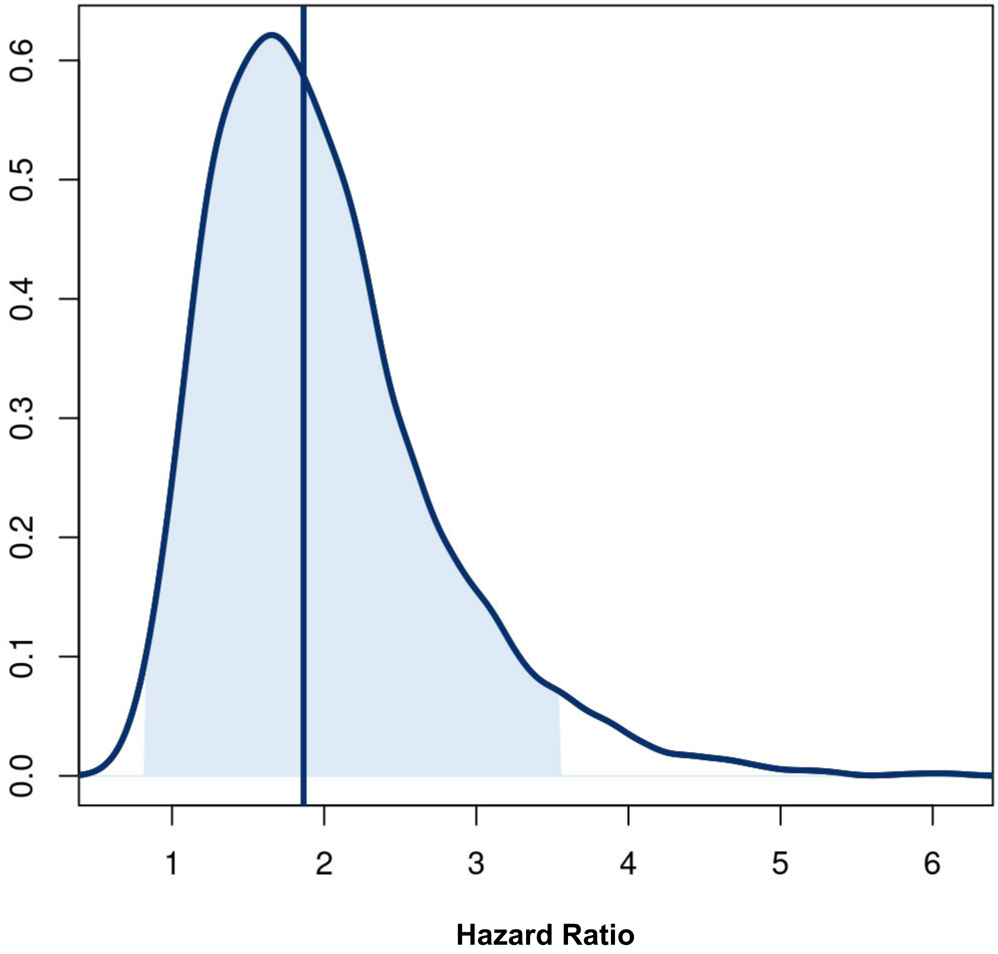

Posterior distribution for the treatment effect hazard ratio for the time to the composite endpoint of hospitalization, urgent care, emergency room visit, or death through day 28. The posterior was based on a covariate adjusted Cox proportional hazards regression with uninformative prior. The baseline hazard was a degree 5 M-spline function. Covariates in the model included age (as restricted cubic spline), sex, duration of symptoms, vaccine status, geographic region, origination from call center, calendar time (as restricted cubic spline), and symptom burden on the day of drug receipt.

Hazard ratios less than one favor the active intervention of fluticasone furoate.

#### Figure S4. Sensitivity analyses for missing daily symptom data on time to sustained recovery

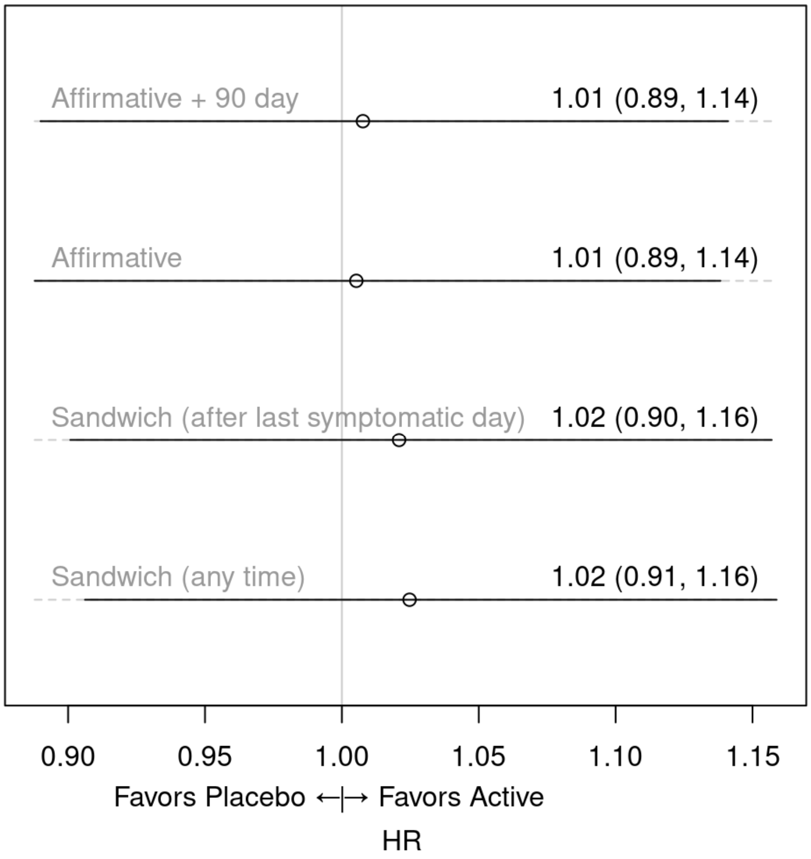

The primary outcome was time to sustained recovery, defined as the third of three consecutive days without symptoms. Participants were queried daily for presence of symptoms and severity. We conducted various sensitivity analyses to assess how missing data may affect the analysis.

- *Affirmative + 90 day:* participant reported three consecutive days of symptom freedom and were symptom free at Day 90. Missed responses coded as symptomatic.
- *Affirmative:* participant reported three consecutive days of symptom freedom. Missed responses coded as symptomatic. This method was used for the primary analysis.
- *Sandwich (after last symptomatic day):* missed survey responses bounded by symptom free days coded as symptom free.
- *Sandwich (any time):* missed survey responses between the last symptomatic day and the first asymptomatic day coded as symptom free.

#### Figure S5. Covid-19 clinical progression scale for inhaled fluticasone vs placebo

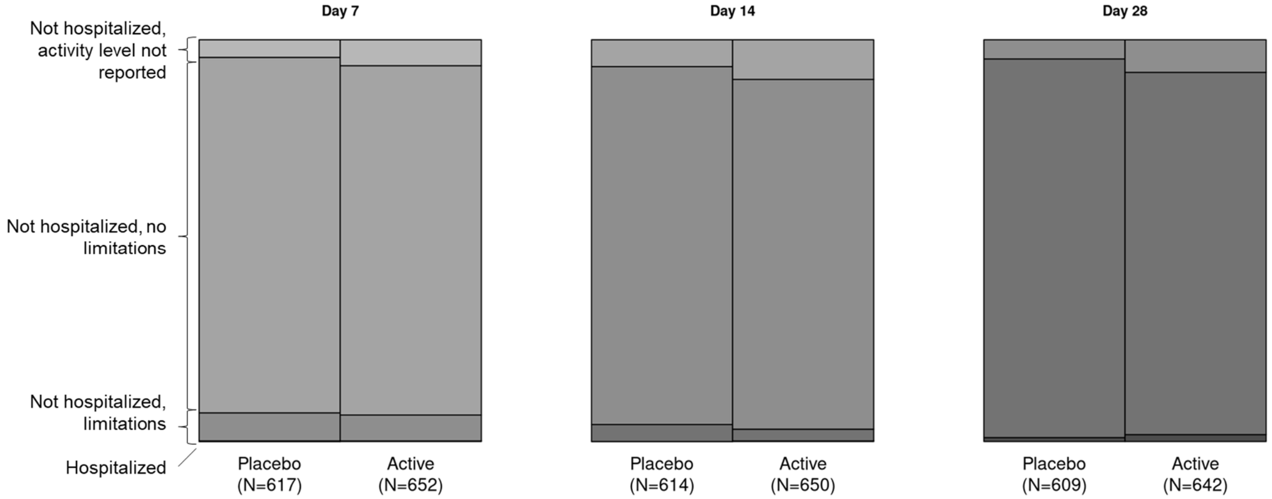

| Day | Day 7 | Day 14 | Day 28 |
|---|---|---|---|
| Odds ratio | 1.28 | 1.33 | 2.06 |
| 95% Credible Interval | 0.85 to 1.70 | 0.88 to 1.84 | 1.21 to 3.05 |
| Posterior P(efficacy) | 0.091 | 0.071 | 0.001 |

Odds ratio >1.0 favors placebo. Refer to Table S3 for numerical data. Posterior P(efficacy) is the probability for efficacy. The Bayesian posterior P (efficacy) is the probability for benefit of the intervention. Thus, the day 28 posterior P(efficacy) = 0.001 implies there is a 0.1% probability that the active arm has a benefit over placebo in the clinical progression scale.
